# Unraveling the metabolomic architecture of autism in a large Danish population-based cohort

**DOI:** 10.1101/2023.11.30.23298767

**Authors:** Filip Ottosson, Francesco Russo, Anna Abrahamsson, Nadia MacSween, Julie Courraud, Kristin Skogstrand, Olle Melander, Ulrika Ericson, Marju Orho-Melander, Arieh S. Cohen, Jakob Grove, Preben Bo Mortensen, David M. Hougaard, Madeleine Ernst

**Affiliations:** Section for Clinical Mass Spectrometry, Danish Center for Neonatal Screening, Department of Congenital Disorders, Statens Serum Institut, Copenhagen, Denmark; iPSYCH, The Lundbeck Foundation Initiative for Integrative Psychiatric Research, Copenhagen, Denmark; Laboratory of Analytical Chemistry, Department of Chemistry, National and Kapodistrian University of Athens, Panepistimiopolis, Zografou, 15771, Athens, Greece; Department of Clinical Therapeutics, School of Medicine, National and Kapodistrian University of Athens, Alexandra Hospital, Athens 11528, Greece; Department of Clinical Sciences, Lund University, Malmö, Sweden; Testcenter Denmark, Statens Serum Institut, Copenhagen, Denmark; Department of Biomedicine - Human Genetics, Aarhus University, Aarhus, Denmark; Bioinformatics Research Center, Aarhus University, Aarhus, Denmark; Center for Genomics and Personalized Medicine, Aarhus, Denmark; NCRR - National Centre for Register-based Research, Aarhus University, Aarhus, Denmark; CIRRAU - Centre for Integrated Registerbased Research at Aarhus University, Aarhus, Denmark

## Abstract

The prevalence of autism in Denmark has been increasing, reaching 1.65% among 10-year-old children and similar trends are seen elsewhere. Although there are several factors associated with autism, including genetic, environmental and prenatal factors, the molecular etiology of autism is largely unknown. Metabolomics has emerged as a tool to measure small molecules that reflect genetic, gut microbiome and dietary intake variations. Here, we apply untargeted metabolomics to over 1400 neonatal dried bloods spots, including neonates who later are diagnosed with autism and matching controls. Overall, we detect underlying molecular perturbations that precede autism related to metabolism of amino acids, acylcarnitines and peptides. In particular the cyclic dipeptide cyclo-leucine-proline and the carnitine-related 5-aminovaleric acid betaine (5-AVAB), were associated with an increased probability for autism. Analysis of genetic and dietary data in over 7000 adults revealed that 5-AVAB was associated with increased habitual dietary intake of dairy and with variants *SLC22A5*, coding for a transmembrane carnitine transporter protein involved in controlling intracellular carnitine levels. We identify 5-AVAB as a novel and potentially modifiable early biomarker for autism that may influence carnitine homeostasis.

## Introduction

Autism refers to a range of neurodevelopmental conditions characterized by impaired social ability, impaired communication and repetitive behavior. The prevalence of autism in Denmark has been increasing, reaching 1.65% among 10-year-old children in 2017^1^. Similar trends are seen elsewhere ^2,3^. Although there are several factors associated with autism, including genetic^4^ and prenatal factors^5^, the molecular etiology of autism is largely unknown. The gut microbiome has also been increasingly implicated in autism, as patients with autism commonly experience gastrointestinal issues^6^ and have an altered gut microbiota^7,8^. Dysbiosis of the maternal gut microbiome could even contribute to altered fetal neurodevelopment related to autism^9^. The circulating metabolome reflects genetic^10^, gut microbiome^11^ and dietary intake variations^12,13^, potentially highlighting mediators between these exposures and autism risk. Detecting autism-related metabolite alterations in early life may facilitate increased understanding of the molecular mechanisms behind autism, improve diagnosis and contribute to preventive strategies.

Metabolomics has been used to characterize specific perturbations of the metabolome in children with autism compared to neurotypical controls, including amino acids^14^, acylcarnitines^15^, and various aromatic and phenolic compounds^15^. However, these studies reflect cross-sectional differences between autism cases and controls, which hampers the attempts to identify early pathophysiological mechanisms of autism, due to the large risk of reverse causation. Metabolites that prospectively associate with autism development have a higher chance of highlighting early protective or detrimental mechanisms of autism that can be targeted in novel preventive strategies.

To unravel underlying disease mechanisms and find tools for early diagnosis of autism, we analyzed the metabolomic profiles of a subset of a large Danish population-based case-cohort sample (iPSYCH2015) consisting of over 1400 newborns, including neonates who later were diagnosed with autism, and matching controls. By using untargeted metabolomics of neonatal dried blood spot samples (DBS), we aim at identifying metabolite features associated with autism, in order to better understand the disease etiology. Metabolites that were associated with autism were further investigated in independent cohorts comprising over 7000 adult individuals, in order to describe the genetic and dietary determinants.

## Results

We utilized a cohort of 1478 male Danish neonates born between 2003 and 2008, comprising 739 newborns who were diagnosed with autism before the age of 10, and 739 typically developed individuals. Since the neonatal dried-blood spot (DBS) metabolome is influenced by the age at sampling (3 to 8 days), season of birth^16^, gestational age^17^ (gestational week 34 to 42) and biobank storage time (15-18 years)^18^, these factors were used to match each case of incident autism with a control neonate. Cases and controls were very similar in terms of gestational age, age at sampling, season of birth and age of mother (Table 1). Family history of mental disorders were significantly more common in incident autism cases compared to controls (p<0.001). The mean age of autism diagnosis was 6.0 years. A total of 865 metabolites (mass spectral features with unique MS/MS fragmentation patterns) were measured and present in at least 25% of the samples in the discovery cohort. Putative annotation on the metabolite class level was conducted by combining mass spectral molecular networking (GNPS)^19^, unsupervised substructure discovery (MS2LDA)^20^, *in silico* annotation through Network Annotation Propagation^21^, Sirius+CSI:FingerID^22^, MolNetEnhancer^23^ and deep neural networks in CANOPUS^24^. This resulted in level 2 annotations for 111 (12.8%) metabolite features and level 3 annotation for 229 (26.5%) metabolite features^25^.

**Table 1.** Characteristics of neonatal study cohort, presented as mean values for continuous variables (standard deviation within parentheses) and percentage for categorical variables (numbers within parentheses).

|  | Cases (N=739) | Controls (N=739) |
| --- | --- | --- |
| **Age at sampling (years)** | 5.8 ($\pm 1.2$) | 5.8 ($\pm 1.1$) |
| **Gestational age (weeks)** | 40.0 ($\pm 1.3$) | 40.0 ($\pm 1.2$) |
| **Born in winter** | 22.7% (168) | 22.7% (168) |
| **Born in spring** | 24.4% (180) | 25.0% (185) |
| **Born in summer** | 25.7% (190) | 25.4% (188) |
| **Born in autumn** | 27.2% (201) | 26.8% (198) |
| **Age of mother (years)** | 30.8 ($\pm 5.1$) | 30.8 ($\pm 4.7$) |
| **Age of autism onset (years)** | 6.0 ($\pm 2.0$) | - |
| **Family history of mental disorder** | 30.0% (222) | 18.7% (138) |

### Metabolites from a wide range of biochemical classes are associated with autism

In total, 99 metabolites had nominally significant associations (p <0.05) with development of autism, as evaluated using logistic regression models adjusted for gestational age, age at sampling, season of birth, and birth year (Figure 1A. Table S1). These included 18 metabolites that could be assigned a structural annotation (level 2 annotation), additionally 15 metabolites with a putative metabolite class and 66 unknown features. The annotated features included several acylcarnitine related metabolites (5-aminovaleric acid betaine, C2:0-carnitine, C8:0-carnitine, C10:0-carnitine, C10:3-carnitine and C16:0-carnitine), cyclic dipeptides (cyclo-Leucine-Proline and cylo-Proline-Valine), tryptophan metabolites (indole-3-acetic acid and indole-carboxaldehyde), methionine cycle metabolites (methionine and betaine), alanine, uric acid, creatine, amino adipic acid, glycerophosphocholine and pantothenic acid.

**Figure 1.**
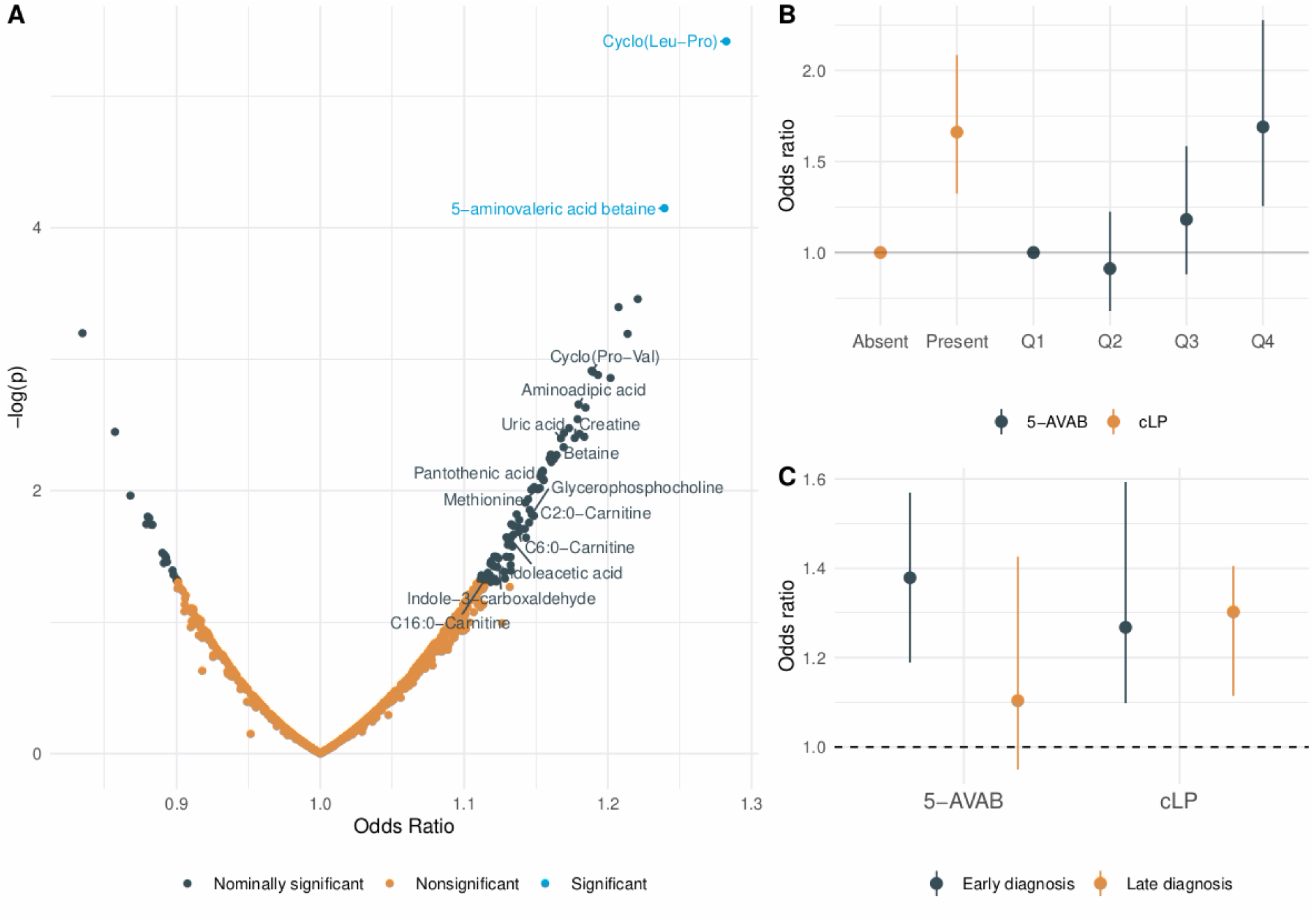
Associations between metabolites in neonatal dried blood spots and future probability of autism spectrum disorder. Odds ratios are calculated using logistic regression models, adjusted for age at sampling, gestational age, birth year and season of birth. **(A)** Associations between all measured metabolite features and autism, with significance threshold defined as false discovery rate adjusted p-value <0.05. Odds ratios are expressed as increased odds of autism per standard deviation increment of metabolite level. Significant features with an annotation confidence of level 2 or higher are named in the figure. **(B)** Associations between 5-aminovaleric acid betaine (5-AVAB) (quartiles) and cyclo-Leucine-Proline (cLP) (binary) and autism. Odds ratios are expressed as compared to the reference category (“Absent” for cLP and quartile 1 [Q1] for 5-AVAB). **(C)** Associations with autism stratified for age of diagnosis (<6 years of age) for 5-AVAB and cLP. Odds ratios are expressed per standard deviation increment of metabolite level.

After adjusting for multiple testing, only two of the 99 metabolites remained significant, cyclo-Leucine-Proline (cLP) (FDR-adjusted p-value=3.3e-3) (Figure S1) and 5-aminovaleric acid betaine (5-AVAB) (FDR-adjusted p-value =0.031) (Figure S2). Being in the top quartile of 5-AVAB was associated with a 69 % increased probability of future autism compared to being in the lowest quartile (Figure 1B). Since measurable levels of cLP were detected in less than 75% of the neonatal cohort (33%), it was treated as a binary variable in the regression analyses. Having measurable levels of cLP was associated with a 70 % increased odds of developing autism in the future (Figure 1B).

Given that autism is partly dependent on genetic factors, we investigated whether the neonatal metabolome was associated with a genetic risk score for autism and with family history of psychiatric disorders. No metabolites were significantly associated (FDR-adjusted p-value <0.05) with the genetic risk score of autism, but 24 nominally significant associations were found (Figure S3). There were eight metabolites associated with family history of psychiatric disorders (FDR-adjusted p-value <0.05), including 5-AVAB, C8:1-carnitine, stachydrine, and trimethyllysine, while a total of 127 metabolites had nominally significant associations (p<0.05) (Figure S4). However, adjustments for family history of mental disorders and genetic risk score for autism only had a minor impact on the association between metabolites and future autism (Table S2).

Overall, the association between the neonatal metabolome and autism was stronger for individuals diagnosed before (N=391), compared to after (N=348) the age of six. There were 40 nominally significant associations for early diagnosed individuals and 13 nominally significant metabolites for individuals diagnosed late. Also, two metabolites, 5-AVAB and an unknown feature (*m/z* 247.13, 3.91 min) remained associated with early diagnosis of autism after adjustments for multiple testing (Table S3). The association between 5-AVAB and autism was significantly stronger (p for interaction = 0.032) in autism cases diagnosed at the age of six years or earlier. For cLP the associations were similar for cases diagnosed before and after the age of six (Figure 1C).

#### Cyclo(leucine-proline) and autism

cLP and the structurally similar cyclo(pro-val) belong to the metabolite class diketopiperazines (cyclic dipeptides), both associated with increased probability of autism in the current study. Both metabolites were detected in less than 75 % of the neonates and were thus treated as binary variables. For cLP, cyclo(pro-val) was the most strongly correlated metabolite, while the caffeine metabolite paraxanthine was the most negatively correlated (Figure 2A). Coffee intake has been positively correlated with plasma cLP levels^26^ and cLP has been found in roasted coffee beans^27^. Moreover, genome-wide association studies (GWAS) have found associations between genetic variants in the *CYP1A2* gene, coding for the rate-limiting enzyme in caffeine metabolism, and plasma levels of cLP ^28^. We did not find any significant correlation between cLP and caffeine in the neonatal DBS (rho=0.00 p=0.97). Moreover, including both cLP and cyclo(pro-val) as covariates in the regression model on autism clearly attenuated the associations for both metabolites (Table S4), indicating that the two diketopiperazines may have a shared pathophysiological relation to autism.

**Figure 2.**
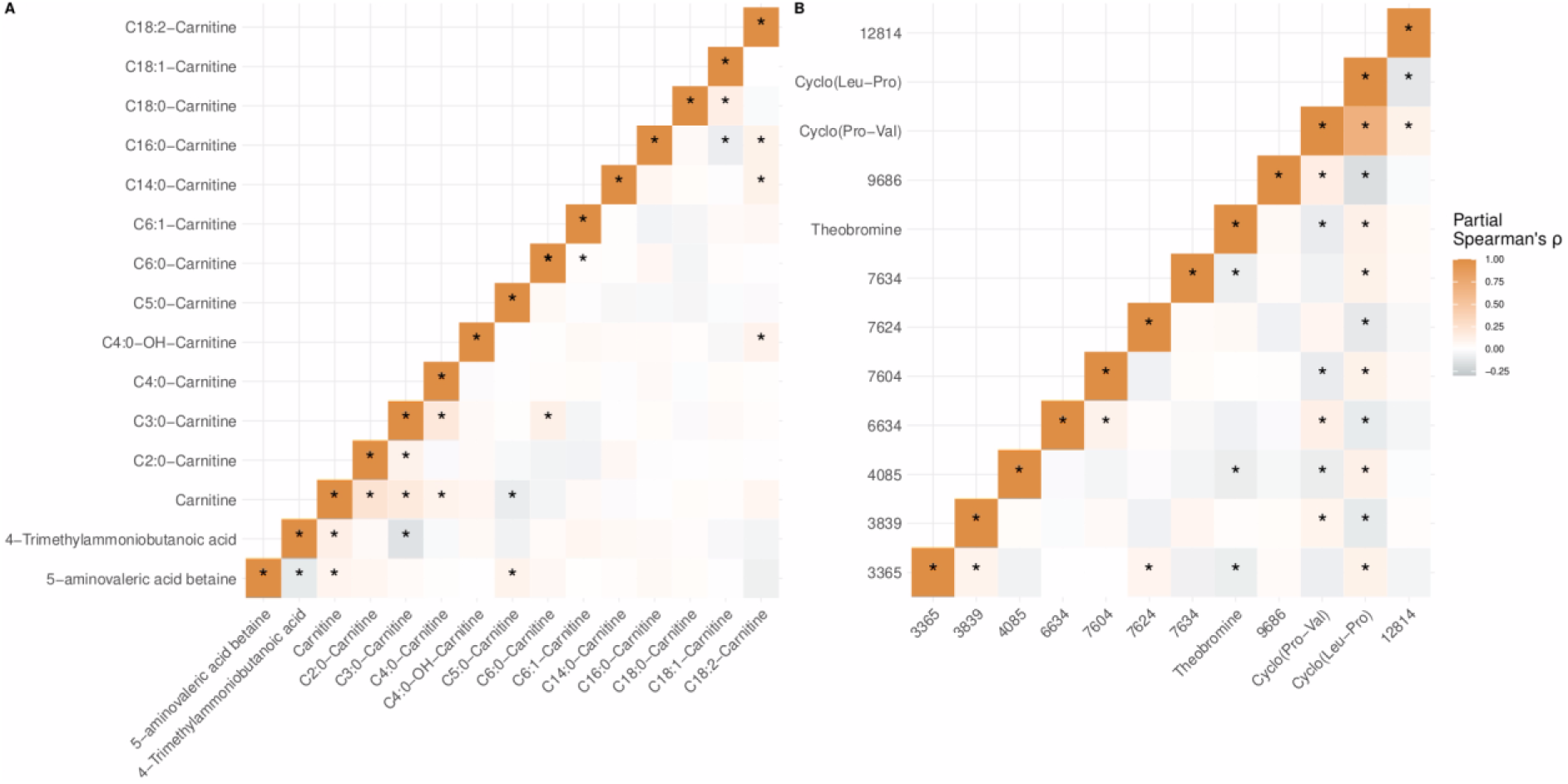
Correlations between the two metabolites significantly associated with autism and other detected metabolites. Correlation coefficients are partial Spearman’s correlation coefficients adjusted for all metabolite features (N=865). Significant correlations are indicated with an * at p<0.05. **(A)** Correlations between acylcarnitine-related metabolites. **(B)** Correlations between cyclo(leucine-proline) and other detected metabolites.

#### 5-aminovaleric acid betaine and autism

5-AVAB is a key regulator of intracellular carnitine availability, since it influences both cellular uptake^29^ and biosynthesis of carnitine^30^. As aforementioned, we also found that elevated levels of several acylcarnitines was associated (p<0.05) with the later development of autism. Metabolite class enrichment analysis showed that acylcarnitines were significantly enriched among nominally significant associations with future autism (p=0.015) and with family history of mental disorders (p=0.013) (Table S5). Partial correlation analysis revealed that 5-AVAB was correlated with several metabolites in the acylcarnitine class (Figure 2B). Notably, 5-AVAB showed positive correlations with C5:0-carnitine and free carnitine but negative correlation with the carnitine precursor 4-trimethylamoniumbutanoic acid. Adjusting the model between 5-AVAB and autism for the autism-associated acylcarnitines (C2:0-carnitine, C8:0-carnitine, C10:0-carnitine, C10:3-carnitine and C16:0-carnitine), the associations were attenuated but remained significant (OR=1.22, P=1.5e-3).

If a causal link between 5-AVAB and autism probability can be established, it is important to examine how neonatal 5-AVAB levels can be modified. We have previously shown that 5-AVAB is vertically transferred from mother to infant, since the maternal levels, both during gestational week 24 and one week postpartum, are strongly correlated with the neonatal levels^31^. This suggests that the neonatal levels of 5-AVAB to a large extent are a reflection of the prenatal exposure of 5-AVAB. Understanding the key determinants of 5-AVAB in adults, should highlight potential strategies to modulate the maternal levels and thus the resulting prenatal exposure.

### Dietary and genetic determinants of 5-aminovaleric acid betaine

To investigate determinants of 5-AVAB levels in adults, we hypothesized that its levels could be influenced by dietary or genetic factors. We used plasma metabolomics, genome-wide genotyping and dietary intake data from two Swedish population-based cohorts, Malmö Diet and Cancer Study (MDC) (N=3833) and Malmö Offspring Study (MOS) (N=3430) (Table S6). A feature with correct MS1 match (*m/z* error < 5 ppm) to 5-AVAB (*m/z* 160.13) was putatively annotated in both cohorts by matching acquired *m/z* 160.13 fragmentation spectra to online databases using MASST^32^. One spectrum achieved a match in both MDC (cosine = 0.83, mass difference <0.01 and 5 shared peaks) and MOS (cosine = 0.81, mass difference <0.01 and 4 shared peaks) with the uploaded 5-AVAB spectrum (Figure S5), and is thus referred to as 5-AVAB from here onwards. In both MDC and MOS, 5-AVAB was correlated similarly with acylcarnitine-related metabolites as in the cohort of neonates. This includes overall positive correlations with short-chain acylcarnitines and free carnitine along with negative correlations with 4-trimethylamoniumbutanoic acid for MOS only (Figure S6-S7).

By investigating six dietary intake groups in MDC (N=3714), 5-AVAB correlated significantly (FDR-adjusted p-value <0.05) with increased intake of dairy (rho=0.18), decreased intake of meat (rho=-0.09), and decreased intake fruit and vegetables (rho=-0.07). Among specific dairy-related intakes, increased intake of milk (rho=0.22) and yogurt (rho=0.09) were significantly correlated (Figure 3A). Similar correlations were seen for 5-AVAB in MOS (N=1539), where dairy intake (rho=0.15) was correlated with 5-AVAB (Figure 3B). Intake of milk (rho=0.12), but not other specific dairy intakes was associated with 5-AVAB in both cohorts. By conducting a genome-wide association study (GWAS) in MDC (N=3409), we found several genome-wide significant (p<5.e-8) variants in the *SLC22A5* and *SLC22A4* genes, with rs272889 being the most strongly associated SNP (p=3.6E-20) (Figure 4)(Table S7).

**Figure 3.**
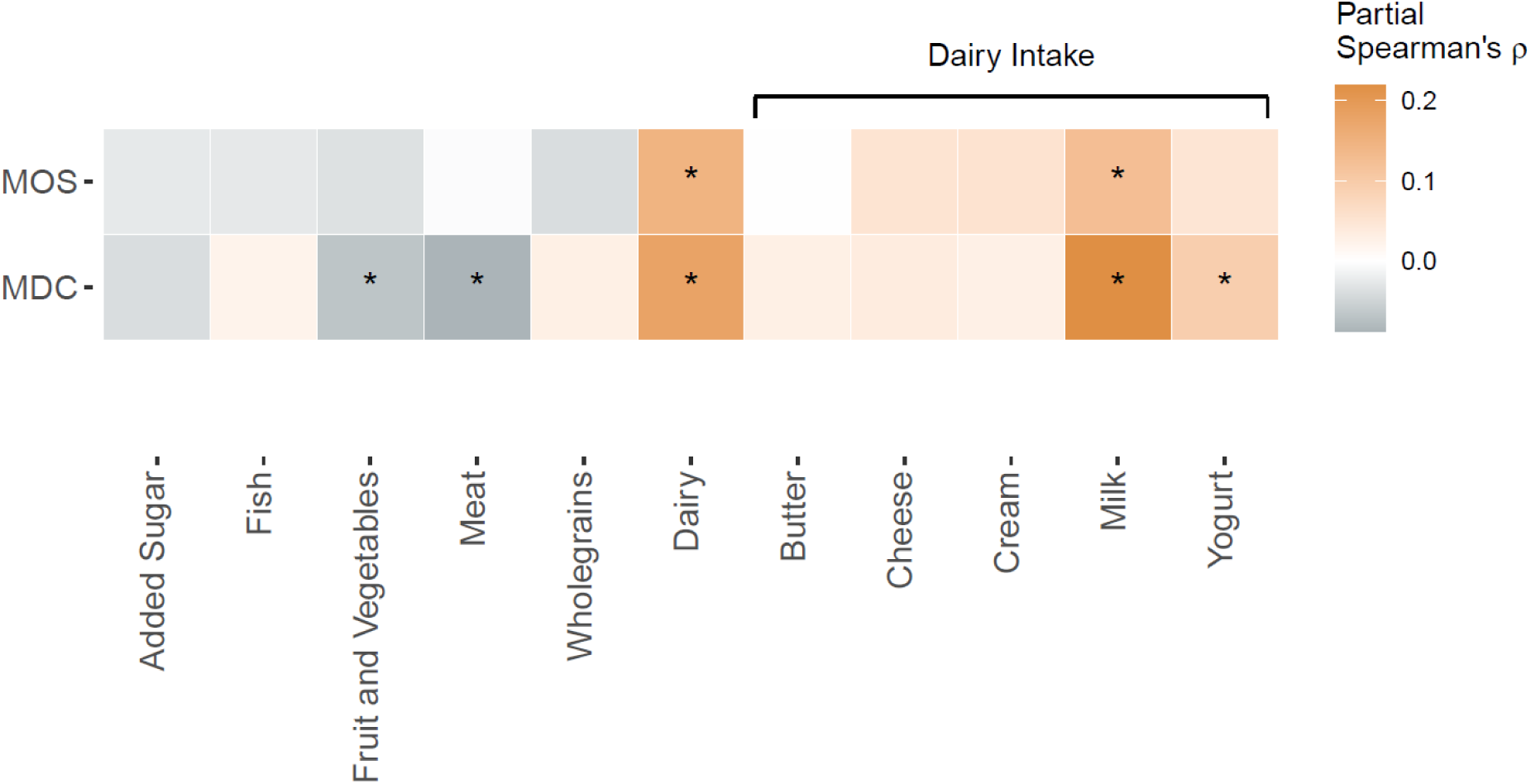
Associations between plasma levels of 5-AVAB and dietary intakes in the Malmö Diet and Cancer Study (N=3714) and the Malmö Offspring Study (N=1539). The heatmap shows partial Spearman’s correlations (adjusted for age, sex and body-mass index) between eleven dietary intake groups and 5-AVAB. Significance was defined as FDR-adjusted p-value <0.05.

**Figure 4.**
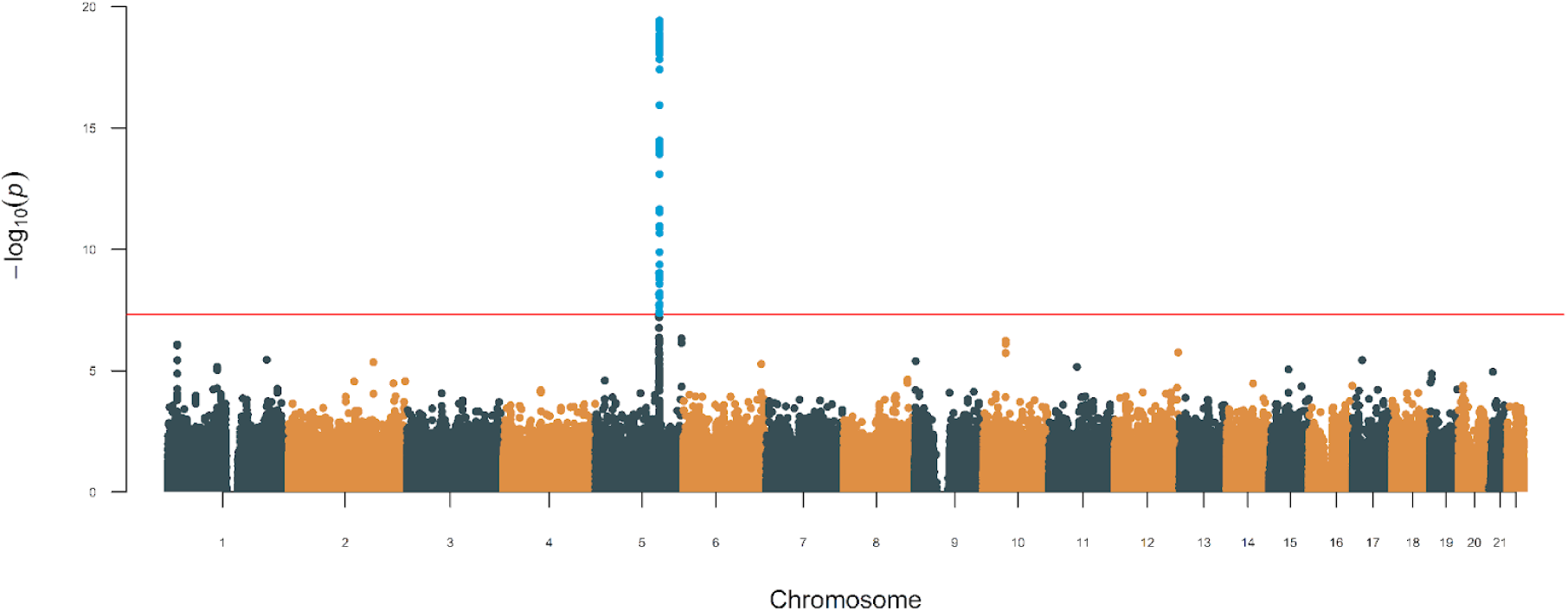
Manhattan plot of the genome-wide association study of plasma levels of 5-aminovaleric acid betaine (5-AVAB) in the Malmö Diet and Cancer Study (MDC) (N=3409). Genome-wide significance threshold is indicated at p<5.0e-8. Genome-wide significant SNPs are colored in blue and are located in the *SLC22A5* and *SLC22A4* genes.

## Discussion

The main finding of our study is that levels of 5-AVAB and cLP are higher in newborns who later develop autism compared to neurotypical controls. We found additional putative associations between metabolites previously linked to autism in cross-sectional studies, indicating that these alterations may be present already at birth. To our knowledge, this is the first large scale study which investigated the untargeted neonatal metabolome in relation to autism, paving the way for the identification of early metabolite biomarker candidates. We suggest that neonatal 5-AVAB levels largely reflect the maternal levels, which may be modified by dietary interventions.

Diketopiperazines, such as cLP, have been proposed to be involved in psychiatric disorders^33^, have an observed ability to cross the blood brain barrier^34^ and are present in a variety of food and beverages^35^. Diet is a potential determinant of cLP, given that its level has been associated with increased intake of coffee^26^ and alcohol^36^ and decreased intake of starchy vegetables ^36^ in previous studies. However, we were not able to identify cLP in the Malmö adult cohorts, where data on nutrition was available, and can therefore only speculate on potential sources of cLP in our study. It is unlikely that maternal coffee consumption represents the major contribution to neonatal cLP, since caffeine was not correlated with cLP in the neonates. Results from GWAS show that genetic variants in *CYP1A2* and *CYP2C19* have been shown to be associated with cLP levels in adults^28^. Our finding that cLP was correlated with the caffeine metabolite paraxanthine, supports that neonatal levels of cLP could be partly regulated by CYP1A2 activity, since it is the enzyme mainly responsible for degradation of caffeine. CYP2C19 is an enzyme involved in the metabolism of several drugs used for psychiatric disorders, including benzodiazepines and selective serotonin receptor inhibitors (SSRI)^37^. It is expressed in the fetal, but not adult, brain and elevated expression has been associated with lower hippocampal volume and increased risk of depression^38^. Prenatal exposure to SSRIs are associated with an increased probability of autism ^39^, although meta-analyses indicate that at least some of the probability increase can be attributed to genetic confounding^40^. Furthermore, cLP could potentially originate from the gut microbiome, as it previously has been proposed to function as a quorum-sensing molecule for certain bacteria, while inhibiting the growth of other bacteria and fungi. Thus, gut microbes might produce cLP to induce growth suppression among competing bacterial species^41^.

The finding that neonatal levels of the metabolite 5-AVAB is associated with increased risk of future development of autism confirms the results from our previous pilot study in 37 neonates and matching controls^16^. Several studies in mice have indicated that 5-AVAB is involved in brain development, both in the fetus and adult mice. For instance, fetuses of germ-free (GF) mice lack several microbially produced metabolites, including 5-AVAB, and simultaneously display a reduced thalamocortical axonogenesis compared to conventionally raised mice. This effect was reversed by injection of 5-AVAB in the GF mice, preserving normal thalamocortical axonogenesis^42^. Given that early brain growth is often faster in children who later develop autism^43^, one could speculate that high fetal brain levels of 5-AVAB may contribute to stimulating brain overgrowth. On the other hand, levels of 5-AVAB have been shown to be increased in the blood and brain of older adults and have negative effects on learning and memory^44^. Overall, these findings indicate that 5-AVAB may be involved in brain development, although the detailed functions are largely unknown.

Speculating that 5-AVAB is related to brain development, it is crucial to investigate how neonatal 5-AVAB levels can be modulated. We have previously shown that 5-AVAB may be directly transferred from the mother to the neonate, as the maternal plasma levels of 5-AVAB is strongly correlated with the neonatal levels of DBS^31^. Thus, it is possible that the neonatal 5-AVAB levels could be considered a proxy for the prenatal exposure to the maternal 5-AVAB.

The current and previous studies indicate that 5-AVAB in adults originates from endogenous production, dietary intake and the gut microbiome, which is briefly discussed below.Endogenous production of 5-AVAB from the carnitine precursor trimethyllysine (TML) has been reported^45,46^ and has been proposed to occur via oxidative deamination followed by decarboxylation^47^. The exact production pathway and to what extent the total pool of circulating 5-AVAB originates from endogenous production is however not known. More evidence for an endogenous production of 5-AVAB comes from a recent study investigating the effects of predicted loss-of-function variants in the X-chromosome gene *TMLHE*^48^. *TMLHE* codes for the enzyme trimethyllysine dioxygenase, responsible for catalyzing the conversion of TML to hydroxytrimethyllysine, the first step in the carnitine biosynthesis. Predicted loss-of-function variants in *TMLHE* associates with increased levels of TML and 5-AVAB and with decreased levels of hydroxytrimethyllysine and 4-trimethylammoniobutanoic, both downstream of *TMLHE* in the carnitine biosynthesis pathway. As expected, the effects of the loss-of-function variants are significantly larger in men compared to women^48^. Although endogenous production is likely an important source of circulating 5-AVAB, there is mounting evidence that the gut microbiome also is a major contributor^29,30^. These include several reports of lack of 5-AVAB in germ-free mice^29,42^, strong associations between microbial species in the human gut and levels of 5-AVAB in the brain ^44^ and plasma^11^ and correlations with gut microbial diversity^11^. Moreover, levels of 5-AVAB in mice fetuses are distinctly diminished when maternal microbiota is lacking^49^. When maternal microbiota is present, 5-AVAB appears to localize to a wide variety of tissues, including the brain^49^. The relative contribution of the endogenous and the gut microbial production of 5-AVAB are not known and interspecies differences may exist. For instance, substantial differences in the biosynthesis of carnitine, where carnitine is produced from TML much more efficiently in mice compared to humans^50^, may increase the bioavailability of TML for production of 5-AVAB in humans. Although the gut microbiota is likely an important source of human circulating 5-AVAB, the exact microbial pathways to produce 5-AVAB are unknown. It has been shown that 5-AVAB can be produced from TML by gut microbes in mice^29^, but is unknown to what extent TML is the source of microbially produced 5-AVAB in humans and other pathways have been proposed ^45^. In recent years, the gut microbiome has increasingly been implicated in autism, as children with autism experience increased prevalence of gastrointestinal issues^6^, have altered gut microbiome^7,8^ compared to typically developing children. Maternal gut health has also been linked to the probability of autism in the offspring^9^, suggesting that the maternal gut microbiome may influence health outcomes for the offspring. In this context, we propose that 5-AVAB may be a mediator in the gut-brain axis. Dietary intake could be an alternative source of 5-AVAB and previous studies have shown that plasma 5-AVAB is correlated with increased intake of whole-grain^51^ and milk^52,53^, which we further support by showing associations between habitual intake of total dairy, milk and yogurt with increased plasma 5-AVAB in two large middle-age Swedish cohorts. No association was seen with whole-grain intake. Although our findings support that dietary intake may be an important contributor to circulating 5-AVAB, we cannot conclude to what extent the contributions from the dietary intake and gut microbiome are independent of each other, since it is possible that specific dietary intake favors 5-AVAB producing gut microbes. Other than endogenous production, the gut microbiome and dietary intakes, genetic factors may also influence the circulating levels of 5-AVAB. We found that plasma 5-AVAB was genome-wide associated with variants in the *SLC22A5* gene, coding for a transmembrane transporter for carnitines and 5-AVAB, supporting results from previous GWAS^54,55^. Overall, our findings indicate that 5-AVAB levels are influenced by both genetic and dietary factors, in addition to the well-established link with the gut microbiome.

The most well described function of 5-AVAB is its ability to influence lipid metabolism via regulating the cellular uptake of carnitine and acylcarnitines. Cellular uptake of 5-AVAB is dependent on the carnitine transport protein *SLC22A5*, whereby it can compete with free carnitine and acylcarnitines for uptake into tissues^29,45,56^ and across the blood-brain barrier ^57^. Moreover, 5-AVAB has been shown to regulate the endogenous synthesis of carnitine in mice by competitive inhibition of *ℽ*-butyrobetaine hydroxylase, the enzyme catalyzing the rate-limiting step in the carnitine biosynthesis pathway, the hydroxylation of 4-Trimethylammoniobutanoic acid (*ℽ*-butyrobetaine) to carnitine^30^. Thus, 5-AVAB regulates carnitine homeostasis by decreasing carnitine biosynthesis and lowering cellular uptake, which in concert may decrease fatty acid oxidation^29,30^ and lead to mitochondrial dysfunction^30^, due to intracellular carnitine deficiency. Our study also indicates an intricate relation between 5-AVAB and acylcarnitines, where levels of 5-AVAB were correlated with higher circulating carnitine and short-chain acylcarnitines but lower levels of 4-trimethylammoniobutanoic acid. Deletions in *TMLHE* are relatively common (1/350 males), cause carriers to be auxotrophic for carnitine and have also been linked to increased probability of developing autism in boys^58,59^. Several previous cross-sectional metabolomics studies show that children with autism have altered circulating levels of acylcarnitines ^15,60,61^ and free carnitine^15^. Carnitine supplementation in children with autism have resulted in improved behavioral scores^62^, but larger studies are needed to confirm their efficacy. The directionality of the associations have not been completely consistent and one study indicates that the acylcarnitines in children with autism differ depending on the fatty acid side chain, where short-chain acylcarnitines generally were more abundant in autism compared to controls and vice versa for long-chain acylcarnitines^15^. In our study this is reflected in correlations between acylcarnitines and 5-AVAB, with a gradient in correlations going from positive for short-chain to negative for long-chain features. However, the distinction between long- and short-chain features were not seen in the association with future autism. This leads us to propose two potential mechanisms linking 5-AVAB to autism. The results from the current studies are consistent with the theory of brain carnitine deficiency as a causal factor for autism and that 5-AVAB may act as regulator of intracellular uptake of carnitine. An alternative hypothesis, where there is a causal link between 5-AVAB and autism, independent of carnitine, should also be further investigated. This could also explain the links between *TMLHE* deletions in males and autism, since *TMLHE* deficiency also results in increased TML and 5-AVAB levels.

If any of the above causal links between prenatal exposure to high 5-AVAB and autism development can be proven, 5-AVAB may represent an excellent early target for autism prevention. This is particularly true if 5-AVAB levels can be modified in a noninvasive manner by dietary interventions (altered dairy intake) or by promoting a more favorable gut microbiome (pro- or prebiotics).

Finally, several of the nominally significant metabolites have previously been linked to autism. Altered products of gut bacterial tryptophan metabolism have previously been reported in children with autism^63^. In our study this was reflected by increased levels of indole-3-acetic acid and indole-3-carboxaldehyde. Uric acid has been reported to be elevated in the urine of children with autism^64^. The methionine cycle, including methionine and betaine, has been suggested to be altered in autism, but conflicting findings make its involvement in autism etiology unclear^65^.

We acknowledge several limitations with our study. Firstly, although the described autism associated alterations in the neonatal metabolome are present shortly after birth and precede autism diagnosis by several years, our study is observational and should be interpreted within that context. It is thus not possible to infer causal relationships between metabolites and development of autism. However, our findings generate hypotheses for mechanisms linking the neonatal metabolites to autism, which should be further investigated in future studies. Secondly, a broader coverage of the metabolome could have been achieved by using a combination of different extraction solvents, chromatographic separation and ionization mode. Since the sample material was limited, we aimed at using a single method with as broad coverage of annotated semi-polar metabolites as possible. Thirdly, we were unable to measure cLP in the independent middle-aged cohort, which limits our potential to investigate its genetic and dietary determinants. Finally, the neonatal cohort consisted of males only, which hinders us from drawing conclusions about the general population.

## Methods

### Cohorts

#### iPSYCH cohorts

The neonatal cohort consisted of individuals who were included in either the population based cohort or as a case in iPSYCH2015^66^, which is a case-control study of psychiatric disorders, nested within the Danish population born between 1981-2008. The neonates included in the current case-control study were born between 2003-2008. All individuals had available information about gestational age, age at sampling and neonatal dried blood spots (DBS) available. Additionally, all samples passed quality control for genotyping. Cases of autism were all male and defined as the first diagnosis of childhood autism (ICD-10: F84.0) in the Danish Psychiatric Central Research Register in 2015 or earlier, at age 1 or older. Controls were selected among males in the population-based cohort in iPSYCH and with none of the following ICD-10 diagnosis codes in the Danish Psychiatric Central Research Register in 2015 or earlier, at age 1 year or older: F84.0, F84.1, F84.2, F84.3, F84.5, F84.8, F84.9, F90.0. Matching of controls was performed based on birth date +/- 14 days, gestational age +/- 4 days and age at sampling +/- 1 day. For neonates born before 2006, autism cases were excluded if diagnosed with mental retardation (F70-F79) before the 10-year birthday in the Danish Psychiatric Central Research Register as April 2017. Seasons are defined as starting from the 1st day of the month: 1st Dec.-28th Feb. is winter, 1st March-31st May is spring, 1st June-31st August is summer, 1st Sept.-30th Nov. is autumn.

#### Malmö Cohorts

The Malmö Diet and Cancer Study is a population-based prospective cohort consisting of 28,449 individuals. The cardiovascular cohort of MDC was designed to study the epidemiology of carotid artery disease. Enrolling participants between 1991 and 1996 ^67^. Citrate plasma was available for 3833 among the 5405 participants with fasted blood samples. Dietary intake data was available for 3714 individuals and genome-wide genotyping for 3409 individuals.

Malmö Offspring Study is an ongoing population-based cohort study where adult (>18 years old) children and grandchildren from the MDC study are recruited^68^. Participants were invited through letter and visited the research clinic where overnight fasting EDTA plasma samples were collected and anthropometric measurements performed. Plasma samples were available for 3430 participants. Dietary intake data was available for 1539 individuals.

### Metabolomics profiling

DBS samples (3.2-mm-diameter punches) were randomly distributed over nineteen 96-well plates (batches). A batch of DBS consisting of adult blood was created before the sample preparation and stored at −20 °C, referred to as external control (EC) samples. Both EC samples and plate specific pools of neonatal samples were analyzed for quality control purposes. Sample preparation was performed by extraction in 80% methanol by being incubated for 45 minutes and subsequent centrifugation. The sample extract (supernatant) was evaporated under nitrogen before being reconstituted in 95 % solvent A (99.8% water and 0.2% formic acid) and 5 % solvent B (49.9% methanol, 49.9% acetonitrile and 0.2% formic acid). Mass spectrometry analysis was performed using a timsTOF Pro mass spectrometer coupled to a UHPLC Elute LC system, Bruker Daltonics (Billerica, MA, US). The analytical separation was performed on an Acquity HSS T3 (100 Å, 2.1 mm x 100 mm, 1.8 µm) column (Waters, Milford, MA, US). The analysis started with 99% solvent A for 1.5 min, thereafter a linear gradient to 95% solvent B during 8.5 min followed by an isocratic condition at 95% mobile phase B for 2.5 min before going back to 99% mobile phase A and equilibration for 2.4 min. Metabolomics preprocessing was done using the Ion Identity Network workflow in MZmine^69,70^ (version 3.3.0). Before statistical analysis, metabolite features present in less than 25% of the samples were removed and features present in fewer than 75% were treated as binary variables (present or absent). This resulted in a final dataset with a total of 865 metabolite features measured, among which 452 features were continuous and 413 were binary variables. Missing values for metabolite features with continuous measurements were further subjected to imputation using missForest ^71^ and subsequent batch correction in WaveICA ^72^. Annotation of metabolite features was performed using mass spectral molecular networking through the GNPS Platform, unsupervised substructure discovery using MS2LDA, *in silico* annotation through Network Annotation Propagation, Sirius+CSI:FingerID, MolNetEnhancer and deep neural networks in CANOPUS. Detailed descriptions of sample preparation, mass spectrometry analysis, preprocessing, annotation and quality control procedures can be found in the supplementary information.

### Relative quantification of 5-aminovaleric acid betaine in Malmö cohorts

Untargeted metabolomics profiles of plasma samples from MOS and MDC were acquired on a UPLC-QTOF (1290 LC, 6550 MS; Agilent Technologies,Santa Clara, CA) as previously described^73^. Relative abundance of 5-aminovaleric acid betaine was retrieved by integrating the peak areas for the *m/z* putatively annotated as 5-AVAB (*m/z* = 160.13) using Agilent Profinder B.06.00 (Agilent Technologies). Normalization was performed using *m/z* 160.13 measurements in repeatedly injected identical pooled quality control (QC) samples. A low-order nonlinear locally estimated smoothing (LOESS) function was fitted to the *m/z* 160.13 areas in the QC samples as a function of the injection order. The *⍶*-parameter (proportion of QC samples used for the correction curve) was set to 2/3. The resulting correction curve was used to normalize the analytical samples as described previously ^74^. MS/MS data was acquired at 20eV and an isolation width of 1.3 *m/z*. Annotation of *m/z* 160.13 was performed using a MASST^32^ search of the fragmentation spectra of *m/z* 160.13, where matching spectra where defined as cosine >0.7, minimum matched peaks=4, parent mass tolerance <0.05 Da and ion tolerance <0.05 Da.

### Genome-wide association study and genetic risk score

Genome-wide genotyping of participants from the MDC and was conducted using the Illumina GSA Bead Chip (Illumina Inc, San Diego, CA). A genome-wide association study (GWAS) for plasma levels of 5-AVAB was performed using a linear additive genetic model adjusting for age and sex, using PLINK v1.9 ^75^.The polygenic score (PRS) is a cross validated internally trained score that was generating by splitting the iPSYCH sample in 50 random, non-overlapping samples of roughly equal size. For each of the 50 subsamples, a GWAS was run on the complement and the sumstats of that was used to train a PRS in the subsample in question. LDAK^76^ was employed to calculate using the SBayesR algorithm^77^ following the best practices of LDAK. The score was subsequently standardized by subset strata. Principal components analysis was performed in PLINK v1.9^75^.

### Dietary intake assessment

Dietary intakes in the MDC were assessed with a method combining a 7-day menu book, a food frequency questionnaire and a 45-minute interview ^78^.

In the MOS dietary intakes for participants was assessed with a web-based 4-day food record, Riksmaten2010, developed by the Swedish National Food Agency ^79^. In both cohorts, six dietary intake groups were energy-adjusted, by dividing intakes with non-alcoholic energy intake.

### Statistical analysis

Associations between metabolite features and autism were analyzed using logistic regression models. Nominally significant associations were defined as P < 0.05, and significant associations as FDR-adjusted P < 0.05. The primary model was adjusted for gestational age, age at sampling,season of birth and birth year. Significant features (FDR-adjusted p-value <0.05) were analyzed as quartiles (5-AVAB) or the secondary model was additionally adjusted for family history of psychiatric disorders, a genetic risk score for autism and six genetic principal components. The analyses were also stratified for age of diagnosis (age of diagnosis <6). Enrichment for metabolite classes were analyzed using Fisher’s exact test. Associations between metabolite features and family history of psychiatric disorders were analyzed using logistic regression models. Corresponding analyses for metabolite features and genetic risk for autism were conducted using linear regression. Analyses for family history of psychiatric disorders were adjusted for gestational age, age at sampling, season of birth and birth year, while analyses on genetic risk score for autism further were adjusted for the first six genetic principal components. Inter-correlation analyses between metabolite features were conducted using Partial Spearman’s correlation tests, adjusted for all metabolite features in each respective dataset. Correlations between metabolites and habitual dietary intakes were conducted using Partial Spearman’s correlation tests, adjusted for age, sex and body-mass index. Analyses using partial correlations were performed in the *ppcor*^80^ package. All statistical analysis was performed in R 4.2.1 and jupyter notebooks are accessible at: https://github.com/ssi-dk/CD-MRG-metabolomics_autism.

### Ethics approval

The neonatal metabolomics study was conducted according to the principles of the Declaration of Helsinki and approved by the Danish Ethics Committee(1-10-72-287-12) and the ethics committee of Lund University approved the study protocols for MOS (DNR 2012/594) and MDC (DNR2009/633).

## Data availability

The data underlying this study are not publicly available due to the Danish Data Protection Act and European Regulation 2016/679 of the European Parliament and of the Council (GDPR) that prohibit distribution of personal data.The data are available from the corresponding authors upon reasonable request and under a data transfer and collaboration agreement.

## Code availability

Source code and jupyter notebooks used for analysis are publicly available at https://github.com/ssi-dk/CD-MRG-metabolomics_autism

## Acknowledgements

This project was funded by the Lundbeck Foundation through The Lundbeck Foundation Initiative for Integrative Psychiatric Research (iPSYCH), grant number R248-2017-2003— Period III: 1 March 2018 to 28 February 2021; R155-2014-1724: Period II: 1 March 2015 to

28 February 2018; R102-A9118: Period I: 1 March 2012 to 28 February 2015. This research has been conducted using the Danish National Biobank resource supported by the Novo Nordisk Foundation. Some of the computing for this project was performed on the GenomeDK cluster. We would like to thank GenomeDK and Aarhus University for providing computational resources and support that contributed to these research results.

## Author Contributions

F.O performed the analysis and visualization. M.E led the investigation. M.E, F.R, D.H, P.B.M, A.S.C, J.C and K.S contributed to neonatal study design. F.O, M.E and F.R drafted the manuscript and analyzed data. F.O, M.E, A.A and N.M contributed to the mass spectrometry methodology of the neonatal dried blood spots. N.M and A.A acquired the neonatal dried blood spot metabolomics data. O.M, M.O.M and U.E contributed to the design of the adult cohort study. F.O contributed to the mass spectrometry methodology, acquisition and analyses of adult plasma samples. F.O and U.E contributed to dietary assessment analyses. J.G provided input to genetic analyses. All authors provided interpretation of the results and critical feedback on the manuscript.

---

**Figure S1.**
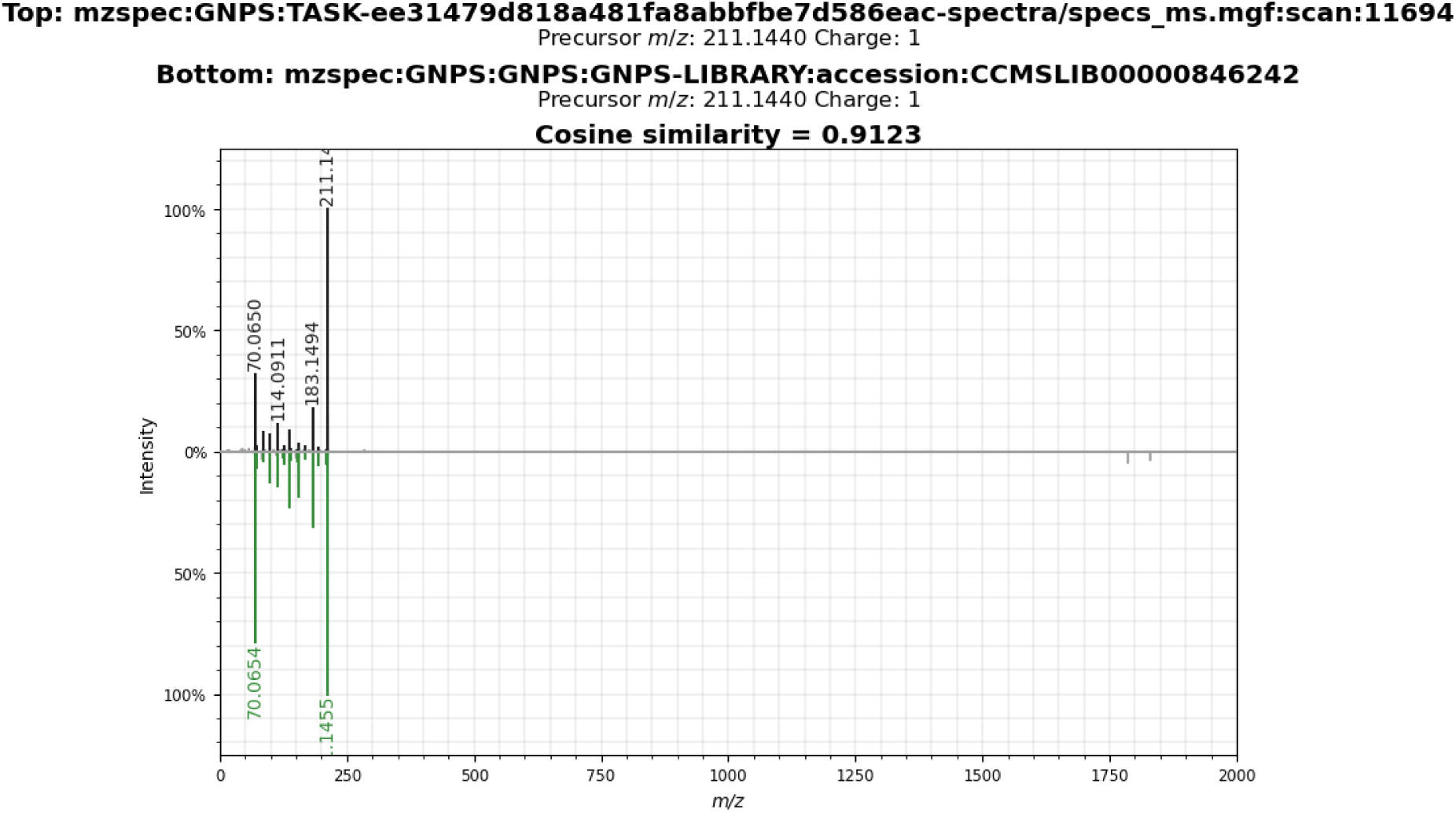
Mirror plot showing the cyclo(leucine-proline) fragmentation spectrum acquired in the neonatal cohort and corresponding library match.

**Figure S2.**
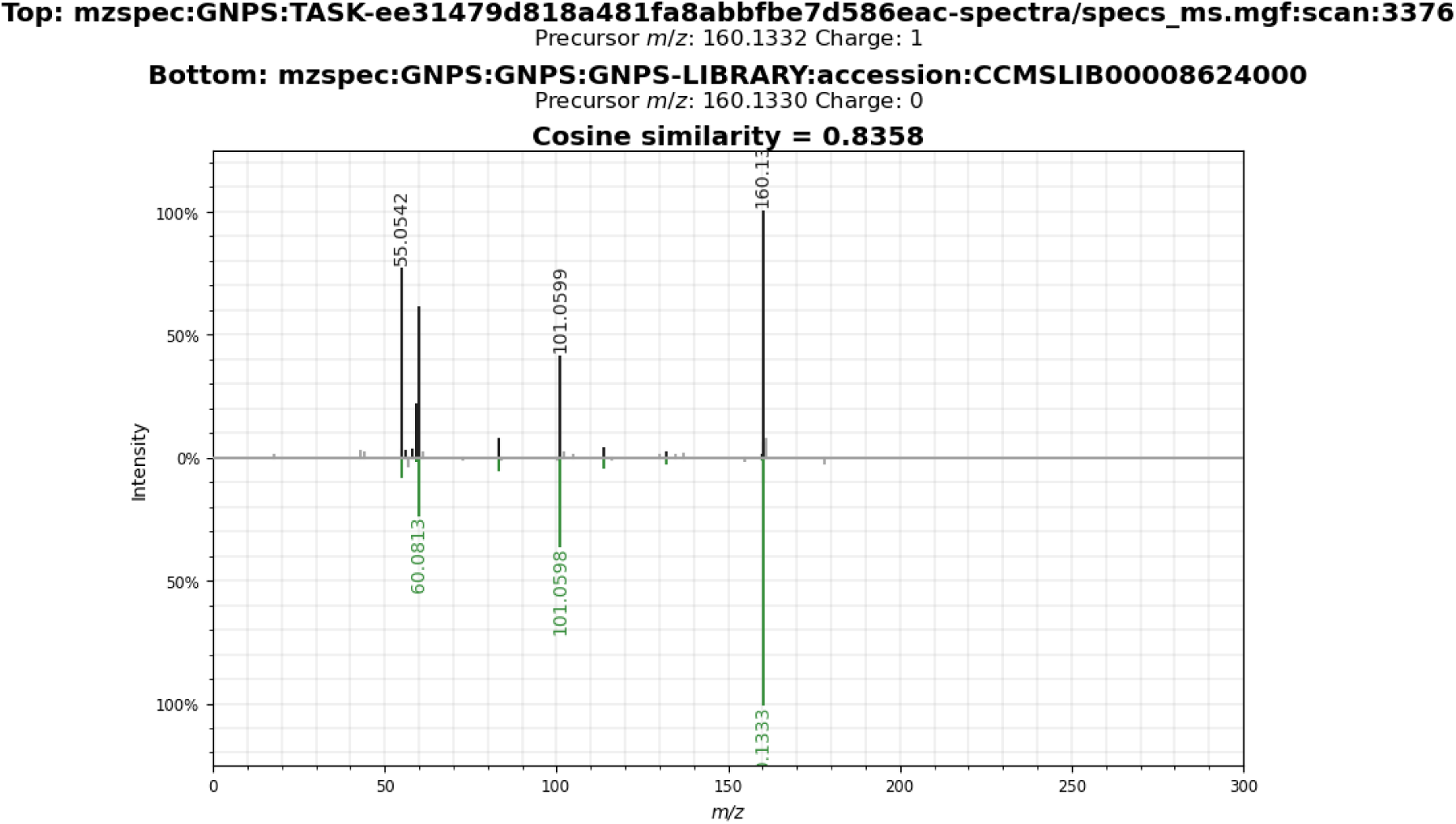
Mirror plot showing the 5-aminovaleric acid betaine fragmentation spectrum acquired in the neonatal cohort and corresponding library match.

**Figure S3.**
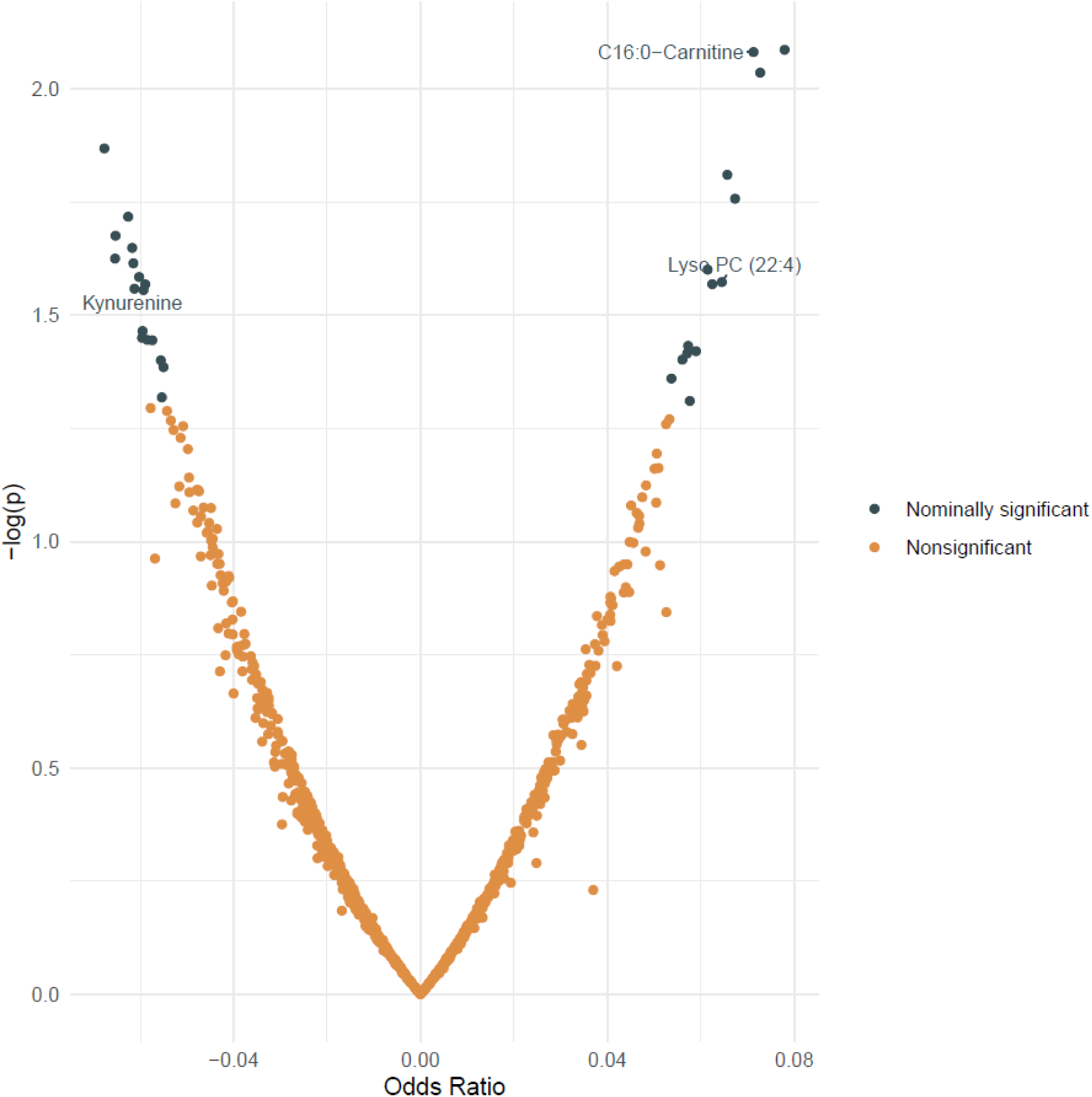
Associations between neonatal metabolites and genetic risk score for autism. Associations were evaluated using logistic regression models adjusted for major factors causing variation in the neonatal dried blood spot metabolome, including gestational age, age at sampling, season and year of birth. Nominally significant associations were defined as P < 0.05, and significant associations as FDR-adjusted P < 0.05.

**Figure S4.**
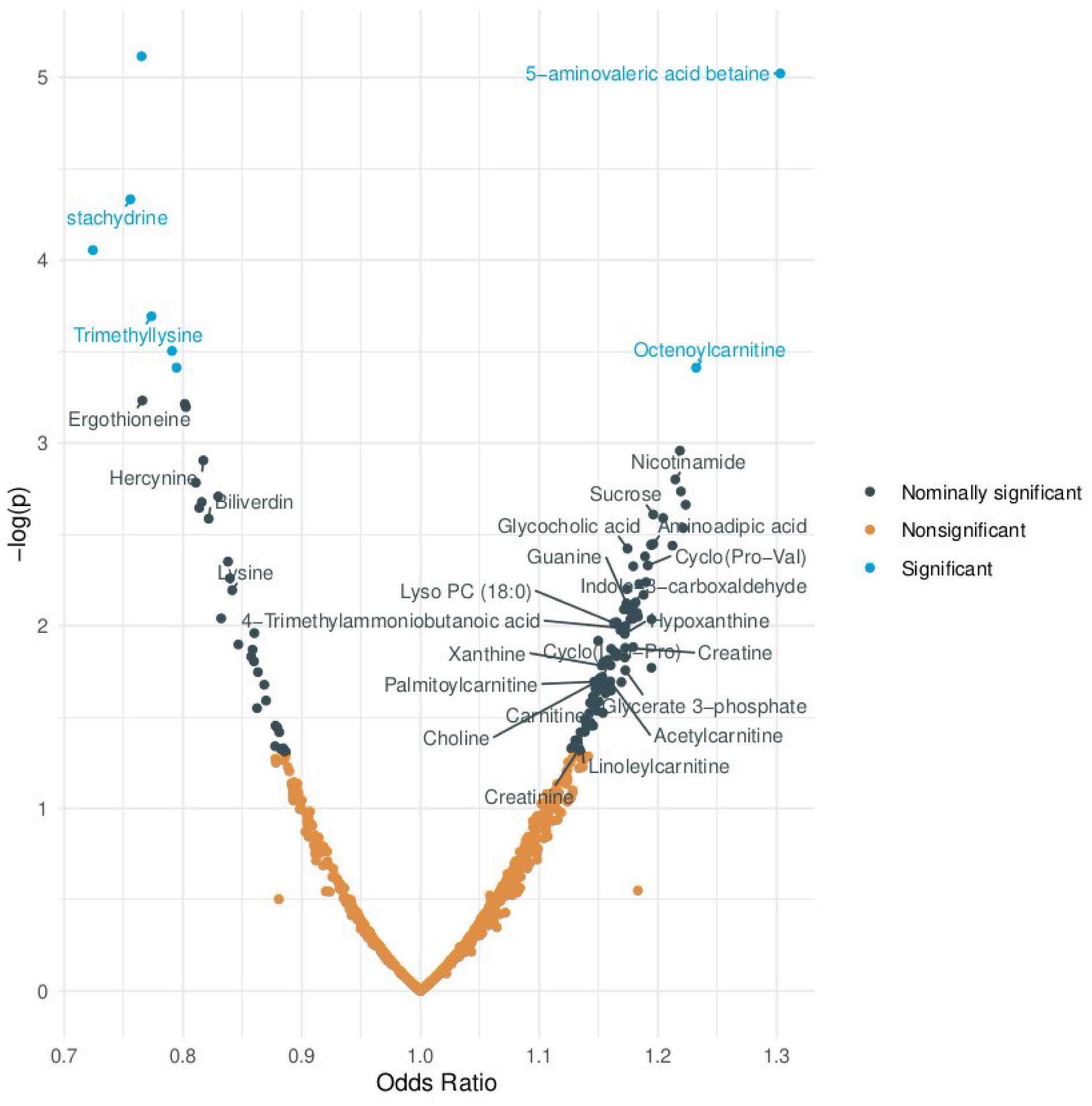
Associations between neonatal metabolite levels and family history of psychiatric disorders. Nominally significant associations were defined as P < 0.05, and significant associations as FDR-adjusted P < 0.05.

**Figure S5.**
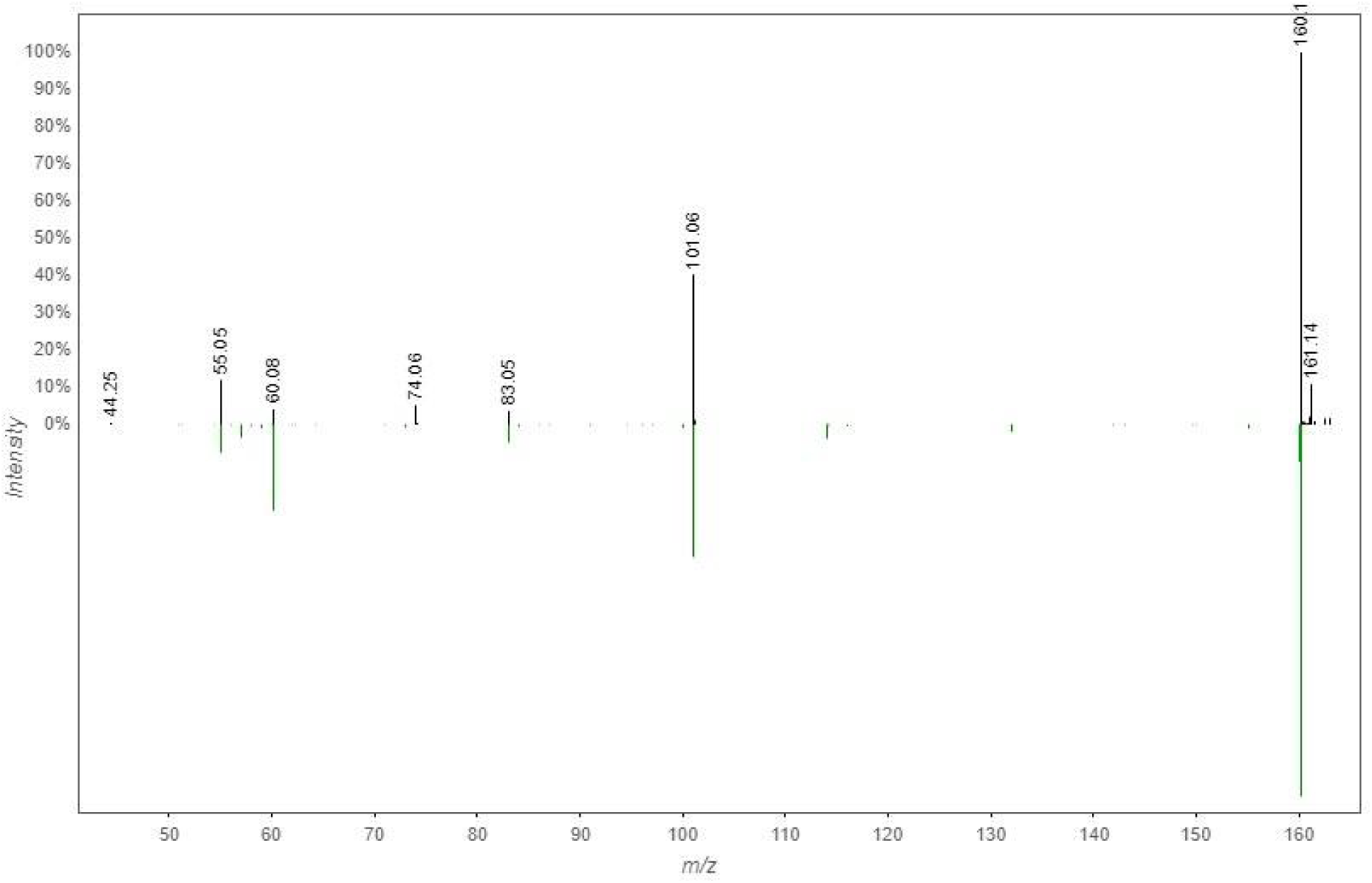
Mirror plot showing the *m/z* 160.13 fragmentation spectrum acquired in the Malmö Offspring Study and corresponding library match to 5-AVAB (cosine=0.81, 4 matching fragments, mass difference <0.01).

**Figure S6.**
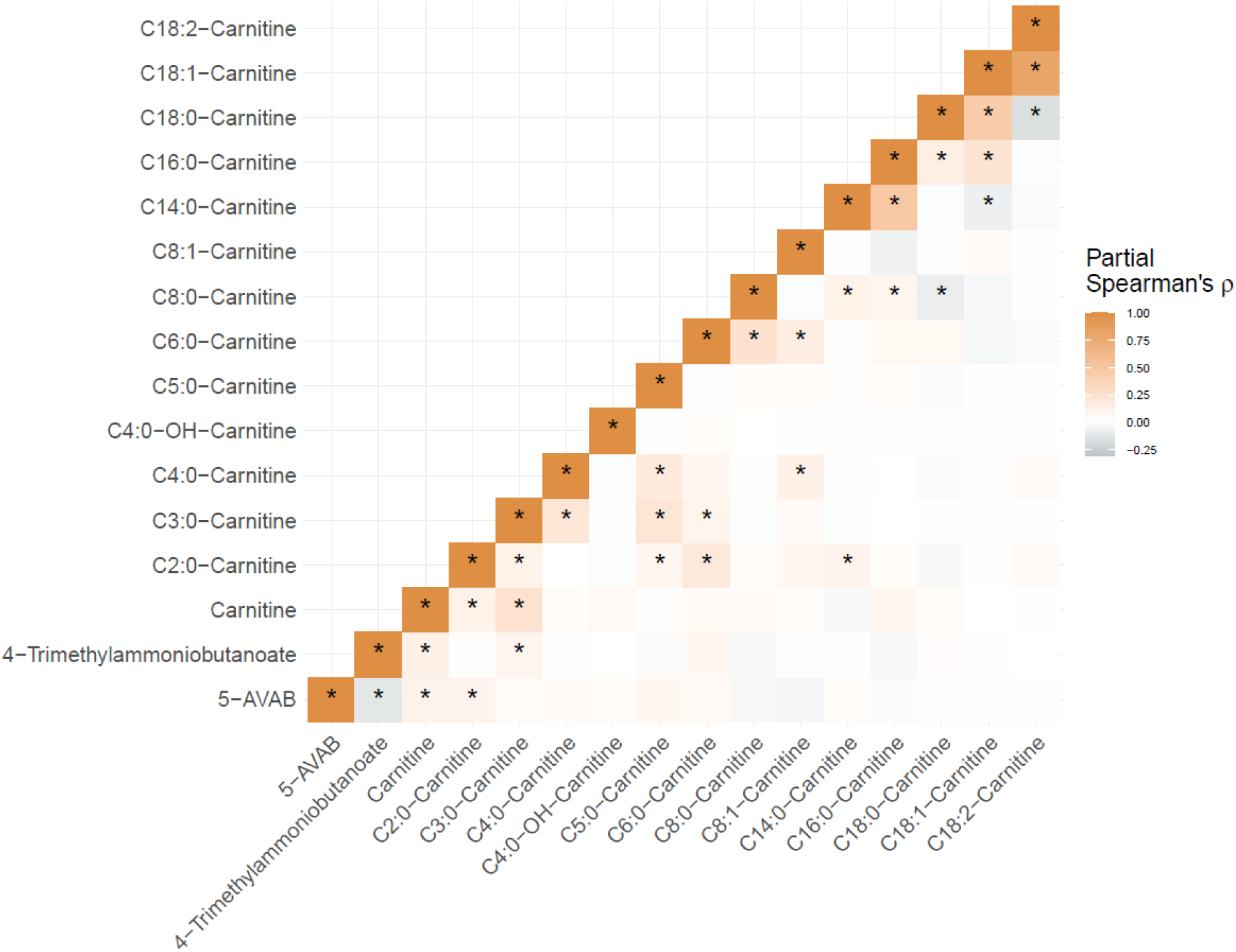
Partial Spearman’s correlation coefficients between carnitine-related metabolites in the Malmö Diet and Cancer Study (N=3833).

**Figure S7.**
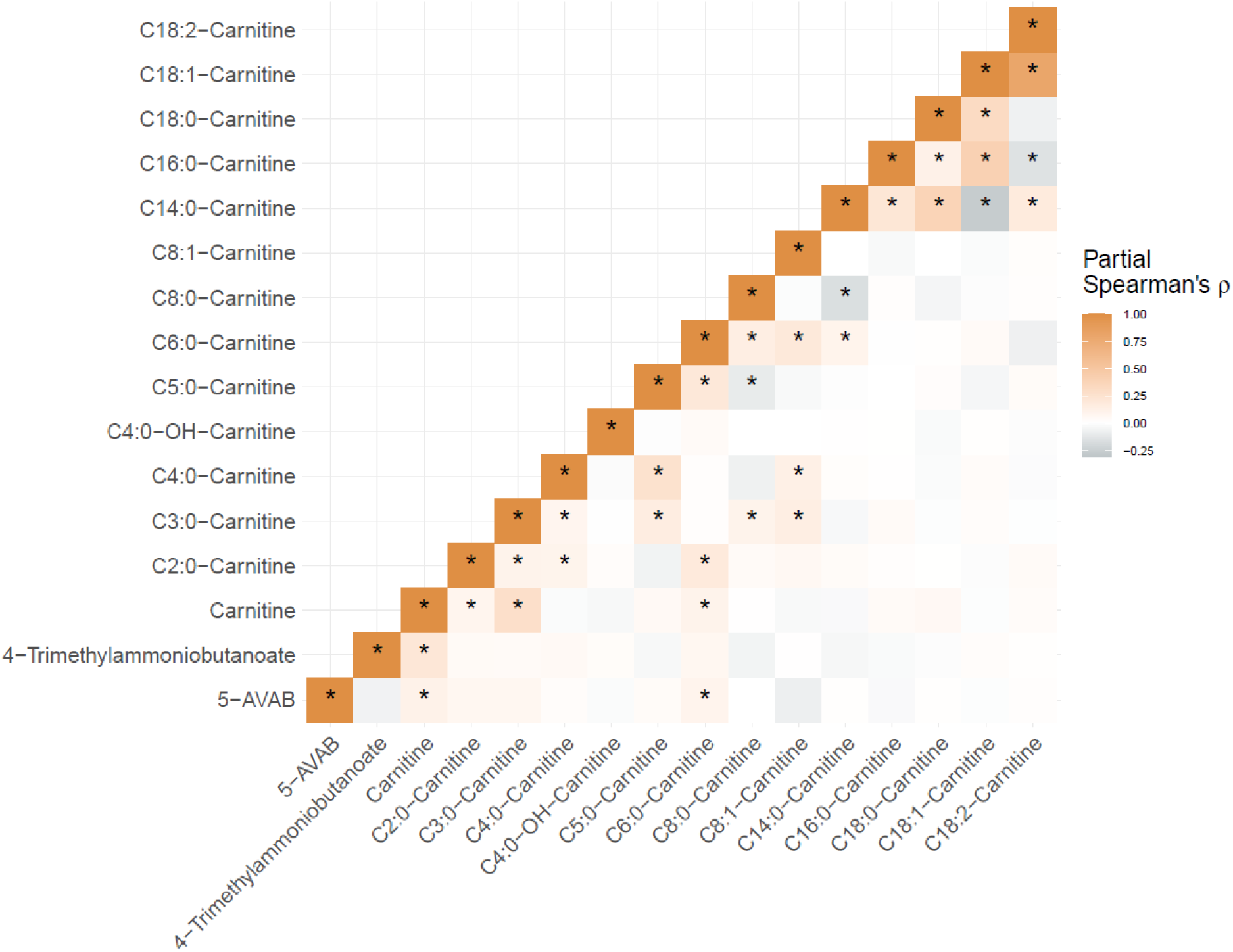
Partial Spearman’s correlation coefficients between carnitine-related metabolites in the Malmö Offspring Study (N=3430).

**Figure S8.**
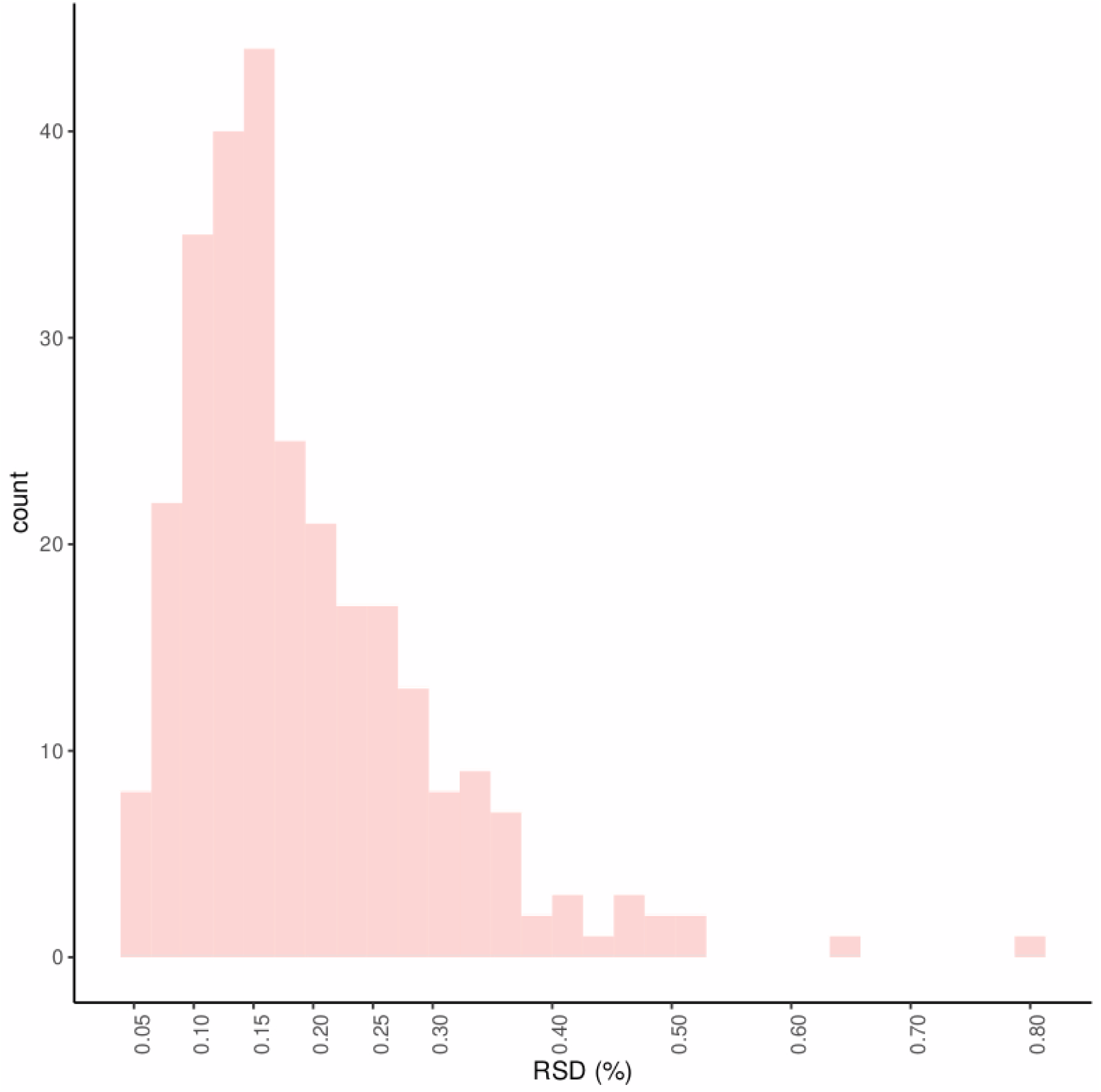
Relative standard deviation (RSD) for metabolite features (N=281) measured in all external control samples.

