## Supplementary information for "Unraveling the metabolomic architecture of autism in a large Danish population-based cohort"

### Sample preparation

Samples were randomly distributed over nineteen 96-well plates (batches). A batch of DBS consisting of adult blood from a single individual was created before the sample preparation and stored at −20 °C. Aliquots (3.2-mm-diameter punches) were distributed on all plates and used as external controls (EC). Plate specific pooled samples were created by aliquoting equal volume of all sample extracts within a plate. Each plate included two water blanks, eight EC, four paper blanks (PB, 3.2-mm diameter punches of blank filter paper), four pooled samples and 80 analytical samples. All solvents were LCMS-grade, and were purchased from Thermo Fisher Scientific (Waltham, MA, USA). DBS samples (3.2-mm-diameter punches) were punched into 96-well plates, made from polypropylene, and kept at −20 °C until extraction. The punching process was done using a Panthera-Puncher 9 from PerkinElmer at room temperature. On the day of extraction, the sample plate was removed from the freezer and kept at room temperature for 30 min. 100 µL of 80% methanol was added to each well, and the plate was then sealed with a silicone plate lid. The plate was then shaken for 45 min at 450 rpm at room temperature, and consecutively centrifuged at 4000 rpm for 30 min at 4 °C. 75 µL of extract was pipetted into a new 96-well polypropylene plate, which was then evaporated under nitrogen for 1 h at 60 L/min, at room temperature. The samples were reconstituted in 75 µL of reconstitution solution (comprised of 5% solvent B in 95% solvent A, see Metabolomics Profiling section), shaken at 600 rpm for 15 min, and then centrifuged at 3000 rpm for 10 min at 4 °C. Afterward the samples on the plate were pooled into a single well on a deep well plate, and pipetted into the four pool positions on the plate, which was then sealed with a silicone lid and centrifuged at 3000 rpm for 5 min at 4 °C. The plate was then run on the LC-MS/MS platform. All pipetting steps were performed on a Microlab STAR automated liquid handler (Hamilton Bonaduz AG, Bonaduz, Switzerland). The extraction procedure took approximately 4.5 h.

### Metabolomics profiling

The LC-MS/MS platform consisted of timsTOF Pro mass spectrometer with an Apollo II ion-source for electrospray ionization, Bruker Daltonics (Billerica, MA, US) coupled to a UHPLC Elute LC system, Bruker Daltonics (Billerica, MA, US). The chromatographic separation system included a binary pump, an autosampler with cooling function, and a column oven with temperature control. For infusion of the reference solution, used for external and internal mass calibration, an additional isocratic pump, Azura Pump P4.1S (Knauer, Berlin, Germany) was used. The analytical separation was performed on an Acquity HSS T3 (100 Å, 2.1 mm x 100 mm, 1.8 µm) column (Waters, Milford, MA, US). The mobile phase consisted of solvent A

(99.8% water and 0.2% formic acid) and B (49.9% methanol, 49.9% acetonitrile and 0.2% formic acid). The analysis started with 99% mobile phase A for 1.5 min, thereafter a linear gradient to 95% mobile phase B during 8.5 min followed by an isocratic condition at 95% mobile phase B for 2.5 min before going back to 99% mobile phase A and equilibration for 2.4 min. Total run time for each injection was 15 min and the analysis time for a full 96-well plate was approximately 25 h. Samples were maintained at +15°C in the autosampler, 5 µL were loaded to the column with a flow rate of 0.4 mL/min and a column temperature of 40 °C.

Tandem mass spectrometric analysis on the timsTOF Pro was performed in the Q-TOF mode with TIMS off, and auto MS/MS on using the following settings: ionization mode set to positive ionization, mass range set to 20 – 1100 *m/z* and a Spectra Rate of 9 Hz (Sample time 0.11s). Source settings as Capillary: 4500 V, Nebulizer Gas: 2.2 Bar, Dry Gas flow: 10 l/min, Dry Gas temperature: 220 °C. Tune settings as follows: Funnel 1 RF and Funnel 2 RF: 200Vpp, isCID: 0 eV, Multipole RF: 60 Vpp, Deflection Delta: 60 V, Quadrupole Ion Energy: 5 eV with a low mass set to 60 *m/z*, Collision Cell Energy set to 7 eV with a pre Pulse Storage of 5 µs. Stepping is used in Basic Mode with a Collison RF from 250 – 750 Vpp, Transfer Time 20 – 50 µs and Timing set to 50% for both. For MS/MS only the collision energy ranges from 100% - 250% with timing set to 50% for both. Auto MS/MS was used with a predefined Cycle Time of 0.5 s, Active Exclusion was used with Exclusion after 3 Spectra and a Release time set to 0.15 min. Dynamic MS/MS spectra acquisition was applied with a target intensity of 20 000 counts, max MS/MS spectra acquisition of 30 Hz (0.03 sec) and min MS/MS spectra acquisition of 16 Hz (0.06 sec). Sodium formate clusters were applied for instrument mass calibration and for internal recalibration of individual samples. A Precursor Exclusion list was used with Exclusion of mass range of 20-60.

### Metabolomics preprocessing

Bruker .d files were exported to the .mzML format using ProteoWizard's MSConvert10 and subsequently preprocessed using the Ion Identity Network workflow in MZmine[1,2] (version 3.3.0). Data was cropped, with chromatogram retention time from 0.4 to 12 min and *m/z* range from 0 to 1100 retained. Then mass lists were created with MS1 intensity above 5E2 and MS2 intensity above 0 retained. The chromatogram was built through the ADAP chromatogram builder by using the following parameters, minimum group size of scans: 5, group intensity threshold: 5E2, minimum highest intensity: 1.5E3, and *m/z* tolerance: 0.002 *m/z* or 5 ppm. The chromatogram was smoothed with a filter width of 5 and further deconvoluted using the MEDIAN *m/z* center calculation, *m/z* range for MS2 scan pairing 0.002 Da and retention time range for MS2 scan pairing 0.3 min. The local minimum search algorithm was used for deconvolution with parameters set to, chromatographic threshold: 85%, minimum RT range (min): 0.01, minimum relative height: 0%, minimum absolute height: 1.5E3, min ratio of peak top/edge: 2, peak duration range (min): 0.01-0.5. The peaks were deisotoped by using the isotopic peak grouper function, with parameters set to, *m/z* tolerance: 0.002 *m/z* or 5 ppm, retention time tolerance: 0.3 min, monotonic shape: on, maximum charge: 2, representative isotope: most intense. Peaks from all samples were aligned, by using the join aligner function with parameters set to, *m/z* tolerance: 0.002 *m/z* or 5 ppm, retention time tolerance: 0.5 min, weight for *m/z*: 75, weight for retention time: 25. Rows were then filtered using the duplicate peak filter with the new average filter mode and *m/z* tolerance set to 0.001 *m/z* or 5 ppm and RT tolerance 0.03 min. Gap-filling was performed using the same mz and RT range gap filler, with a *m/z* tolerance of 0.002 *m/z* or 5ppm and a RT tolerance of 0.03 minutes. The metaCorrelate function was used to find correlating peak shapes with parameters set to, RT tolerance: 0.1 min, min height: 1E3, noise level: 5E2, min samples in all: 2 (abs), min samples in group: 0 (abs), min %-intensity overlap: 60%, exclude estimated features (gap-filled): on. Parameters for the correlation grouping were set as follows, min data points: 5, min data points on edge: 2, measure: Pearson, min feature shape correlation: 85%. Ion identity networking parameters were set to, *m/z* tolerance: 0.002 *m/z* or 5 ppm, check: one feature, min height: 1E3 with ion identity library parameters set to, MS mode: positive, maximum charge: 2, maximum molecules/cluster: 2, adducts: M+H, M+Na, M+K, modifications: M-H2O, M-NH3. Further ion identity networks were added with *m/z* tolerance: 0.002 *m/z* or 5 ppm, min height: 1E3 and ion identity library parameters set to, MS mode: positive, maximum charge: 2, maximum molecules/cluster: 6, adducts: M+H, M+Na, modifications: M-H2O, M-2H2O, M-3H2O, M-4H2O, M-5H2O and *m/z* tolerance: 0.002 *m/z* or 5 ppm, min height: 1E3, and annotation refinement on with parameters set to, delete smaller networks: link threshold: 4, delete networks without monomer: on, and ion identity library parameters set to MS mode: positive, maximum charge: 2, maximum molecules/cluster: 2, adducts: M+H, M+Na, M+K, modifications: M-H2O, M-NH3. Finally, two feature tables were exported in the .csv format. One feature table containing all extracted mass spectral features and another feature table filtered for mass spectral features with associated fragmentation spectra (MS2). An aggregated list of MS2 fragmentation spectra was exported in the .mgf format and submitted to ion identity feature-based mass spectral molecular networking through the Global Natural Products Social Molecular Networking Platform (GNPS)[3,4].

Before statistical analysis, connected ion adducts were merged and mass spectral feature signals with a relative intensity less than 5 times the mean relative intensity in all paper blank samples were removed. Metabolite features present in less than 25% of the samples were removed and features present in fewer than 75% were treated as binary variables (present or absent). This resulted in a final dataset with a total of 865 metabolite features measured, among which 452 features were continuous and 413 were binary variables. Missing values for metabolite features with continuous measurements were further subjected to imputation and batch correction procedures. Among the 452 metabolite features, 274 (61%) had less than 5% missing values. Missing values were imputed using missForest [5], with the maximum number of iterations set to 10 and the number of trees to 100. Batch correction was performed using WaveICA [6], with the “Haar” wavelet function, maximum components set to 20 and the batch threshold to 0.25.

### Quality control procedures

Quality control procedures for the metabolite profiling are divided into three main categories, system suitability test (SST), batch evaluation and post-processing quality control.

In the SST, the mass spectral and chromatographic performance was evaluated prior to each batch by injecting two different standard samples. Standard sample A consisted of leucine enkephalin (1.8 µM in 50/50: H2O/ACN) and standard sample B consisted of a mix of amino acids and acylcarnitines in 50/50: H2O/ACN (Cambridge Isotope Laboratories, Tewksbury, MA, USA). System suitability was evaluated based on retention time deviation (<0.2 min), mass accuracy (<2 ppm) and relative standard deviation (<20%) for all compounds in both standard sample A and B. Batch evaluation was performed by monitoring sixteen quality control metabolites in pooled sample extracts, EC samples and paper blanks. Potential carry-over is controlled by ensuring that quality control metabolites are not present in the paper blank samples. Mass spectral and chromatographic performance is evaluated by monitoring retention time deviation (<0.2 min), mass accuracy (<2 ppm) and coefficient of variation (<20%) in EC samples and pooled sample extracts. Data for batch evaluation is presented in Table S8. Feature picking for the SST and batch evaluation was performed in Metaboscape (Bruker, Billerica, MA, United States). Post-processing quality control was performed for all features that were detected in all EC samples (N=281), by calculating relative standard deviations (RSD) for each feature. In total, the average RSD was 19 % and 88 % of the features (N=246) had an RSD<30% (Figure S8).

### Metabolite identification

To annotate mass spectral features to putative chemical structures, a mass spectral molecular network was created through the GNPS Platform (http://gnps.ucsd.edu) using the ion identity feature based molecular networking workflow (https://ccms-ucsd.github.io/GNPSDocumentation/fbmn-iin/)[1,3,4]. The data was filtered by removing all MS/MS fragment ions within +/- 17 Da of the precursor *m/z*. MS/MS spectra were window filtered by choosing only the top 6 fragment ions in the +/- 50 Da window throughout the spectrum. The precursor ion mass tolerance was set to 0.02 Da and a MS/MS fragment ion tolerance of 0.02 Da. A network was then created where edges were filtered to have a cosine score above 0.7 and more than 4 matched peaks. Further, edges between two nodes were kept in the network if and only if each of the nodes appeared in each other's respective top 10 most similar nodes. Finally, the maximum size of a molecular family was set to 100, and the lowest scoring edges were removed from molecular families until the molecular family size was below this threshold. The spectra in the network were then searched against all GNPS' spectral libraries. The library spectra were filtered in the same manner as the input data. All matches kept between network spectra and library spectra were required to have a score above 0.7 and at least 4 matched peaks.

To further enhance chemical structural information within the molecular network, substructure in- formation was incorporated into the network using the GNPS MS2LDA workflow (https://ccms-ucsd.github.io/GNPSDocumentation/ms2lda/)[7–9]. Furthermore, information from *in silico* structure annotations from Network Annotation Propagation[10] and Sirius+CSI:FingerID[11] were incorporated into the network using the GNPS MolNetEnhancer workflow (https://ccms-ucsd.github.io/GNPSDocumentation/molnetenhancer/)[12]. Chemical class annotations were performed using deep neural networks in CANOPUS[13] and followed the ClassyFire chemical ontology[14].
